# Effects of D-Glucosamine on the Growth of Human Gut-Dominant Microbiota *In Vitro* and Bowel Movements in Healthy Individuals

**DOI:** 10.1101/2025.06.15.25329540

**Authors:** Tomoya Shintani, Soyoka Sakiyama, Yuta Ami, Hideya Shintani, Shin Kurihara

## Abstract

D-Glucosamine (GlcN), a monomer produced by the hydrolysis of chitosan, is a dietary supplement used worldwide to mitigate cartilage degeneration. Previous reports have shown that some dietary glucosamine migrates to the colon. However, the effect of glucosamine alone on colonic microbiota and bowel movements remains poorly understood. In this study, we evaluated the effect of glucosamine on the growth of 46 dominant human colonic bacterial species and 24 other important bacteria in vitro. Among the 70 gut bacterial species tested, the growth of 57 (81%) was significantly enhanced by 0.5% (w/v) GlcN, with the most prominent growth activity (>5-fold) observed in *Anaerotruncus colihominis*, *Pseudoflavonifractor capillosus*, and *Roseburia hominis*. These results indicate that a wide range of the tested gut bacteria can utilize GlcN, similar to the effect of conventional dietary fiber in improving bowel function. Next, we conducted an open-label, single-arm trial involving 29 healthy individuals to determine the effects of 1500 mg GlcN/day, a commonly used dose. Stool color significantly changed during the 2 weeks of GlcN intake from brown to ocher (*p* < 0.01), suggesting enhanced colonic fermentation. The stool odor and the sensation of incomplete evacuation improved significantly (*p* < 0.05). Numerical measurements of bowel movements revealed significant increases in stool volume, defecation frequency, and the number of days of defecation during GlcN intake (*p* < 0.001). Thus, dietary glucosamine may stimulate gut microbiota growth in the colon and promote bowel movements. This study was registered with the University Hospital Medical Information Network (regd. no. UMIN000056757).

## INTRODUCTION

D-Glucosamine (2-amino-2-deoxy-D-glucose, GlcN) is a fundamental component of chitosan and chitin, which are naturally produced by arthropods [1]. Industrially, GlcN produced as a dietary supplement by hydrolyzing crustacean exoskeletons, which are primarily composed of chitin [2]. GlcN is commonly consumed worldwide as a supplement to alleviate symptoms of osteoarthritis and joint degeneration. However, a meta-analysis has found that GlcN may not be effective in treating joint disorders [3]. Therefore, the anti-osteoarthritis benefits of GlcN remain controversial. Despite this, many people continue to use GlcN as a dietary supplement. A large-scale epidemiological study of supplement users found that GlcN consumption was associated with reduced overall mortality [4,5]. The results of this study are consistent with findings from animal experiments in mice and nematodes [6,7]. Nevertheless, the molecular mechanisms behind GlcN’s potential benefits on longevity remain unknown.

After ingestion, GlcN undergoes partial absorption in the small intestine, with a significant portion remaining unmetabolized and reaching the large intestine [8]. Once in the colon, GlcN may interact with gut microbiota, potentially serving as a substrate for microbial fermentation [9]. This process has been suggested to influence the composition and metabolic activity of intestinal microbiota, leading to the production of short-chain fatty acids (SCFAs). SCFAs play crucial roles in maintaining the intestinal barrier integrity, modulating immune responses, supporting overall metabolic health, and promoting bowel movements [10]. For this reason, it is possible that GlcN contributes to reduced mortality through the intestinal microbiota [11]; however, the details remain unknown.

Several studies have reported that GlcN supplementation may improve bowel movements, further linking its effects to gut microbiota modulation [12]. However, a common limitation of these reports is the relatively small number of study participants, despite significant differences in the intestinal microbiome of each participant, which could restrict the generalizability of the findings. In this study, we cultured and used the most dominant gut commensal microbial species from the large populations of Japan [13] and Europe [14], whose strains could be cultured simultaneously in vitro. We first evaluated the effects of GlcN on the growth of 70 human-dominant colonic bacterial species and other important human bacteria in vitro. Furthermore, we conducted an open-label single-arm trial on the effects of GlcN on bowel improvement in a sufficient number of healthy individuals, more than double the number of participants previously reported.

## MATERIALS AND METHODS

### In vitro assay for growth of human gut bacterium

1. ***Chemicals.*** GlcN hydrochloride was purchased from Fujifilm-Wako (Tokyo, Japan). Other chemicals (not mentioned in the following sections) were obtained from Wako Pure Chemical Industries (Osaka, Japan).
2. ***Microbe strains.*** Bacteria were obtained from the American Type Culture Collection (ATCC), German Collection of Microorganisms and Cell Cultures GmbH (DSMZ), and Japan Collection of Microorganisms (JCM; **Table 1**) [15,16]. Bacteria were cultured at 37 °C in an anaerobic chamber (10% CO_2_, 10% H_2_, and 80% N_2_; InvivO_2_ 400; Ruskinn Technology, Bridgend, UK).
3. ***Microbe medium.*** The procedure for the preparation of GB medium (GAM and blood medium) has been reported in detail previously [15]. Briefly, materials other than blood were autoclaved, placed in a closed container together with Aneropack Kenki (Mitsubishi Gas Chemical Company, Tokyo, Japan), and allowed to stand overnight to remove dissolved oxygen. Horse blood (horse whole-blood, defibrinated and sterile; Nippon Bio-Supp. Center, Tokyo, Japan) stored anaerobically with Aneropack Kenki was added to the Gifu anaerobic medium (GAM) at 5% (v/v) in an anaerobic chamber. GAM supplemented with blood medium (GB) was prepared. GAM (Nissui Pharmaceutical, Tokyo, Japan) was autoclaved (115 °C, 15 min), immediately placed in a closed container together with Aneropack Kenki (Mitsubishi Gas Chemical Company, Tokyo, Japan), and allowed to stand overnight to remove dissolved oxygen. Horse blood that was stored anaerobically in Aneropack Kenki (Mitsubishi Gas Chemical Company, Tokyo, Japan) was then added to GAM at 5% (v/v) in an anaerobic chamber. Semisolid GAM without dextrose broth (Nissui Pharmaceutical) was dissolved following the manufacturer’s instructions and filtered to remove agar [17]. After autoclave sterilization, the medium was immediately placed in a closed container with Aneropack Kenki and allowed to stand overnight to remove oxygen.
4. ***Microbe culture.*** Bacteria were cultured at 37 °C in an anaerobic chamber (10% CO_2_, 10% H_2_, and 80% N_2_; InvivO_2_ 400; Ruskinn Technology, Bridgend, UK). First, 5 μL of thawed frozen cells of the bacterial strains in glycerol at –80 °C was added to 500 μL of GB medium in a deep 96-well plate to inoculate each species, and pre-culture was performed at 37 °C under anaerobic conditions. For pre-culturing in vials, 500 µL of the pre-culture solution was transferred to a deep 96-well plate before using a copy stand. Approximately 2 µL of the respective culture collection was inoculated in 500 μL of GAM medium or GAM medium supplemented with 0.5% GlcN in another deep 96-well plate using a copy plate stand (Tokken, Chiba, Japan). After 48 h of anaerobic incubation, growth was measured at an optical density of 600 nm (OD_600_) using a Thermo Scientific™ Multiskan™ GO instrument (Thermo Fisher Scientific, Waltham, MA).
5. ***Evaluation of GlcN on the growth of gut microbiome in vitro*.** The growth of the most dominant and important intestinal bacteria in the intestinal microbiota of Westerners and Japanese people cultured in a sugar-free medium supplemented with GlcN was substituted to calculate the growth promotion and inhibition effects of GlcN on intestinal bacterial species [18]. Briefly, after 48 h of anaerobic incubation, growth was measured at an optical density of 600 nm (OD_600_). To compare the ability of the bacterial species to utilize GlcN, the OD_600_ ratio was obtained by dividing the bacterial growth (OD_600_) in the medium supplemented with GlcN by that of the medium without GlcN. All outcomes were presented as mean ± SD. Statistical analyses were performed using a two-sided Student’s t-test, and the statistical significance was set at 5 %. The software used was Microsoft Excel 2010 (Microsoft Co., Ltd., Tokyo, Japan).

### Single arm trial on bowel movement in healthy individuals

1. ***Study design, ethics, and participants.*** This was an open-label, single-arm trial. The Shiba Palace Clinic Institutional Review Board approved the study protocol on January 16, 2025 (approval no. 155708_te-37811). The study was conducted with full consideration of medical ethics and in accordance with the Declaration of Helsinki (2013) [19] and the Ethical Guidelines for Medical and Health Research Involving Human Subjects. Testing was conducted by SOUKEN Co., Ltd. This study was registered with the University Hospital Medical Information Network (regd. no. UMIN000056757).
2. ***Screening of participants.*** A follow-up flowchart of the open-label, single-arm trial is shown in Figure 1. The study participants were publicly recruited. A total of forty-nine individuals who agreed to participate were selected from a public database. A preliminary questionnaire was administered to those who provided written informed consent confirming their wish to participate in the study. Constipation-prone individuals aged 20–59 years with a mean frequency of bowel movements of about 3–5 times per week were enrolled in the study. Among them, those who did not meet the following 9 exclusion criteria were selected for participation in the study: 1) Those taking medicines that may affect the test results (medicines with intestinal regulating effects, antibiotics, etc.); 2) Those who regularly consumed health foods that may affect the test results (lactic acid bacteria, oligosaccharides, supplements that claim to regulate the intestines, etc.) or health foods that contain the same main ingredients as the test product; 3) Pregnant or potentially pregnant and breastfeeding women; 4) Those with alcoholism; 5) Those who may have allergic reactions to the test product ingredients; 6) Those participating in other clinical trials; 7) Those with a history of severe liver damage, kidney damage, or heart disease; 8) Those with a history of hepatitis or current illness; and 9) Those with severe anemia. After screening 40 participants, 29 underwent an intake test. The mean age of the 29 participants (22 women, 7 men) was 42.9 ± 9.8 years (mean age: women = 42.9 ± 9.9 years, men = 43.3 ± 10.4 years).
3. ***Test food and intake.*** The test food consisted of a tablet containing GlcN (Houkouen Seiyaku Co., Ltd., Kagawa, Japan). The tablets contained 1500 mg GlcN, 312 mg starch, 300 mg cellulose, 240 mg sucrose fatty acid ester, and 48 mg silicon dioxide. The participants ingested the GlcN (1500 mg) tablets each day with normal or warm water. The study schedules included a no-intake period (2 weeks) and an intake period (2 weeks). All 29 participants completed the study. Twenty-four participants had 100% compliance; the remaining five had 92% compliance. Data from all 29 participants were included in the final analysis.
4. ***Bowel movement questionnaire.*** Participants maintained a bowel diary throughout the 4-week study to record bowel movements and assess defecation and constipation symptoms. The bowel diary recorded the presence and status of bowel movements, including both defecation and constipation-related items. The procedure is described in detail elsewhere [20] (Fig. 1).
5. ***Defecation items assessed on a scale.*** The bowel movement diary of the four defecation items (stool characteristics, color, odor, and residual stool sensation) was assessed on a scale for each item. The stool characteristics were scored using the Bristol stool scale [21]. It classified the stool form into seven scales: 1) “Separate hard lumps, like nuts,” 2) “Sausage-shaped, but lumpy,” 3) “Like a sausage but with cracks on its surface,” 4) “Like a sausage or snake, smooth and soft,” 5) “Soft blobs with clear cut edges,” 6) “Fluffy pieces with ragged edges, a mushy stool,” and 7) “Watery, no solid pieces, entirely liquid.” Each participant selected a score corresponding to the stool, and the scores for each answer were tallied. The closer it was to 4, the more likely it was for the patient to have a better bowel movement status. Stool color was assessed on a 6-point scale (1 = yellow, 2 = light ocher, 3 = ocher, 4 = brown, 5 = dark brown, and 6 = dark brown almost black), the scores for each answer were tallied. Stool odor intensity was assessed on a 5-point scale (1 = very weak, 2 = weak, 3 = normal, 4 = strong, and 5 = very strong), the scores for each answer were tallied. Residual Stool Sensation was assessed on a 4-point scale (1 = no sensation, 2 = almost no sensation, 3 = a little sensation, and 4 = full sensation). The higher the scores for color, odor, and residual stool sensation, the more likely it was for the patient to have a worse bowel movement status. The score of all 4 items (stool characteristics, color, odor, and residual stool sensation) was assessed as the score for 2 weeks (average of the 2-week period).
6. ***Constipation items assessed on a value.*** The bowel movement diary of the four items (amount of stool, frequency of defecation and flatulence, and days of defecation) was subjectively assessed for each item, and the higher the score, the less likely it was to be constipation. The amount of stool per a week was assessed as the estimated number (average of the 2-week period) of chicken eggs (large) converted from the amount of stool. The frequency of defecation (average of the 2-week period) was assessed as the number of defecation events per a week. The frequency of flatulence per a week was assessed as the number (average of the 2-week period) of flatulence events in the tested 2 weeks. The number of defecation days was assessed as the number (average/week) of days for defecation events per a week.
7. ***Statistical Analysis.*** All outcomes were presented as median values and mean ± SD. All items were examined using participant testing in the pre-ingestion-2 week and post-ingestion-2 week periods. All statistical analyses were performed using two-sided testing, and the statistical significance was set at *p* < 0.05. For the scores of stool characteristics, color, odor, and residual stool sensation, the Wilcoxon signed-rank test was performed. For the amount of stool, frequency of defecation and flatulence, and days of defecation, a paired Student’s t-test was performed. The GraphPad Prism software (USACO Co., Ltd., Tokyo, Japan) was used for all the analyses.

## RESULTS

### Effects on gut microbe growth by GlcN

The growth values of the most dominant and important intestinal bacteria in the intestinal microbiota of Westerners and Japanese people, which were cultured in GAM medium and sugar-free GAM medium supplemented with 0.5% (w/v) GlcN, were calculated. To investigate the effect of existing prebiotics on the growth of human gut microbiota, we cultured beneficial, pathogenic, and prominent bacteria (Table 1) in GAM without sugar (GAM-wos) supplemented with GlcN. The prominent bacteria used in this test included 46 dominant species that could be cultured in GAM-wos medium. However, *Faecalibacterium duncaniae* was excluded because its growth was unstable in GAM-wos.

**Fig. 1.**
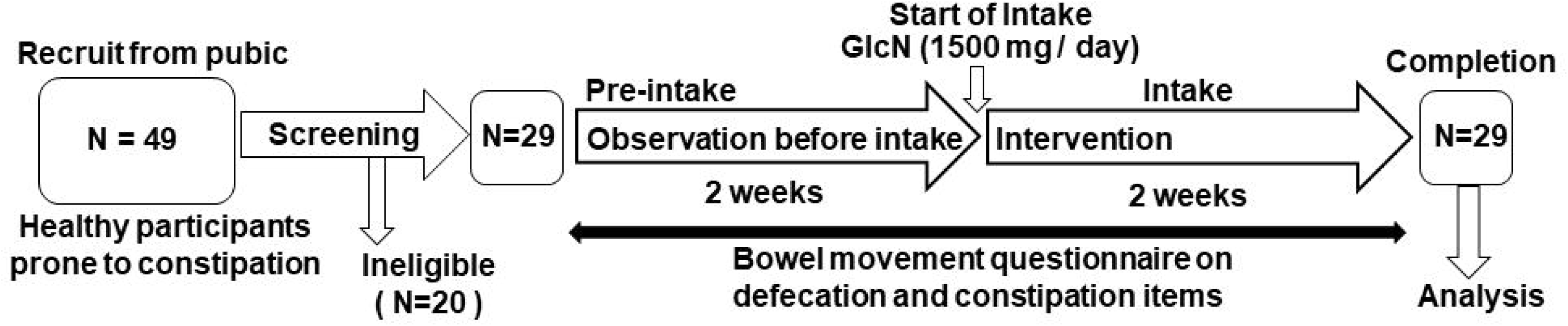
The follow-up flow chart of the open label single arm trial. The study participants were publicly recruited. Forty-nine individuals who agreed to participate were selected from a public database. A preliminary questionnaire was administered to those who provided written informed consent confirming their wish to participate in the study. Constipation-prone persons aged 20–59 years or younger with a mean frequency of bowel movements of about 3–5 times per week were enrolled in the study. Those who did not meet the exclusion criteria were included in the study.

**Table 1.**
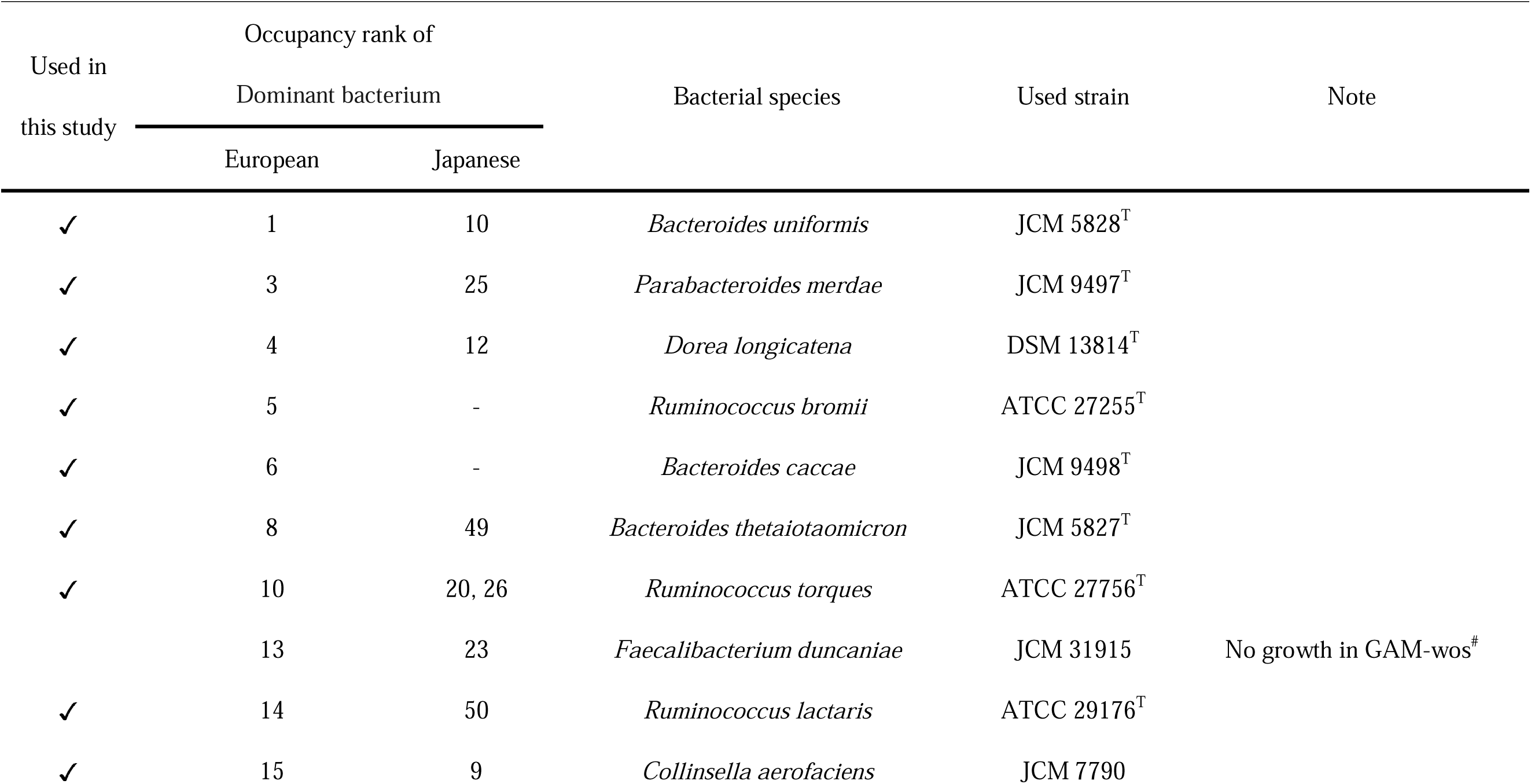
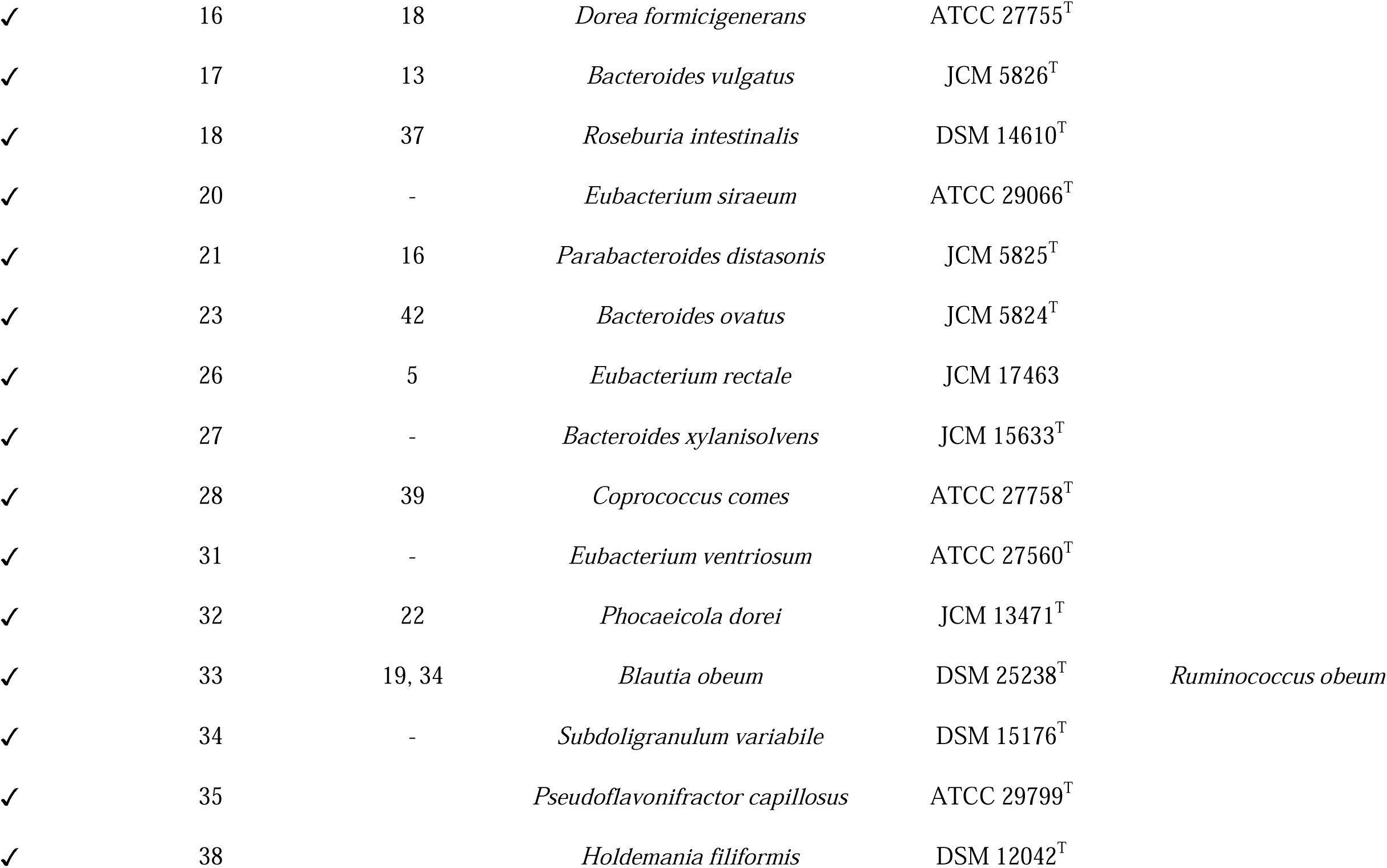
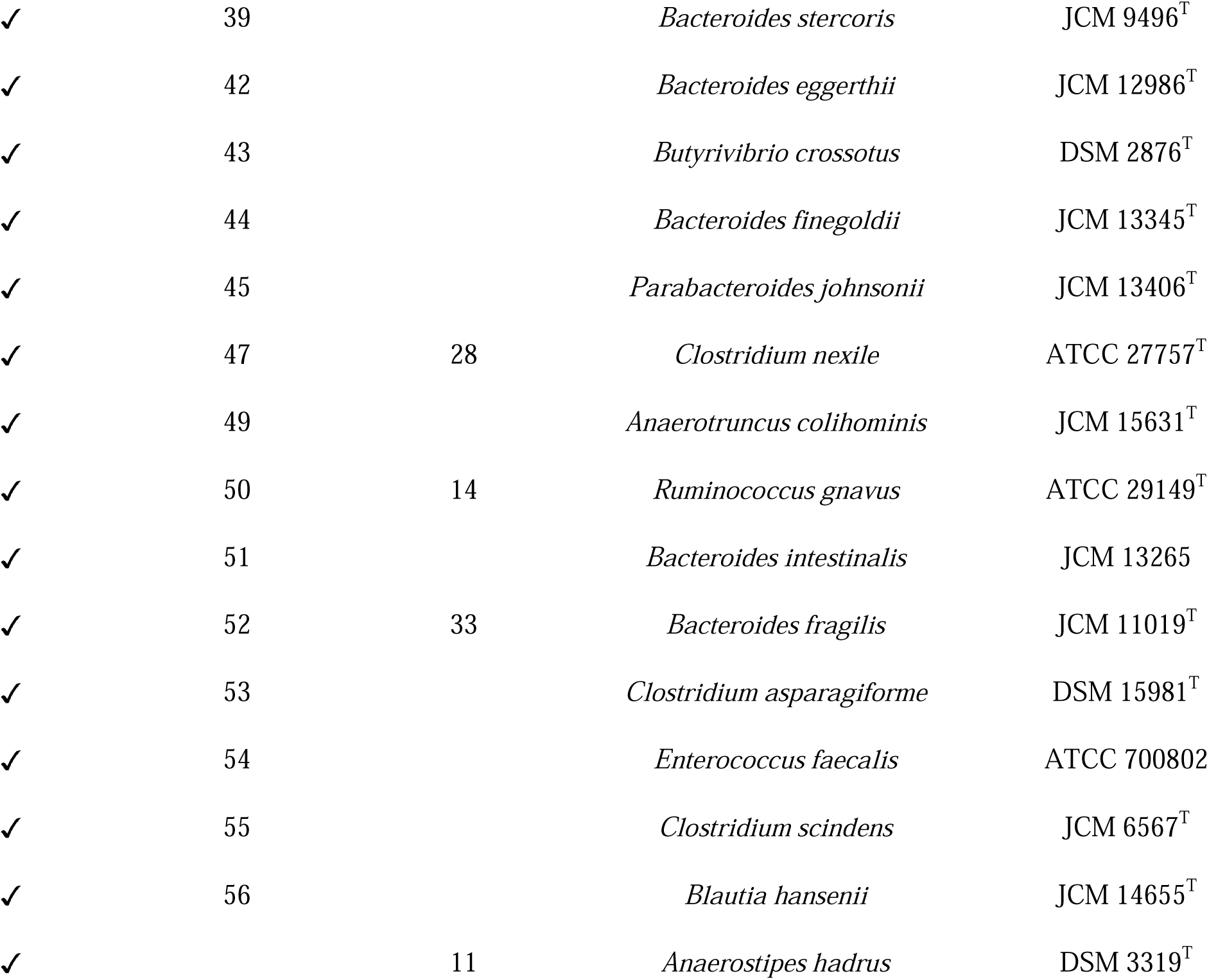
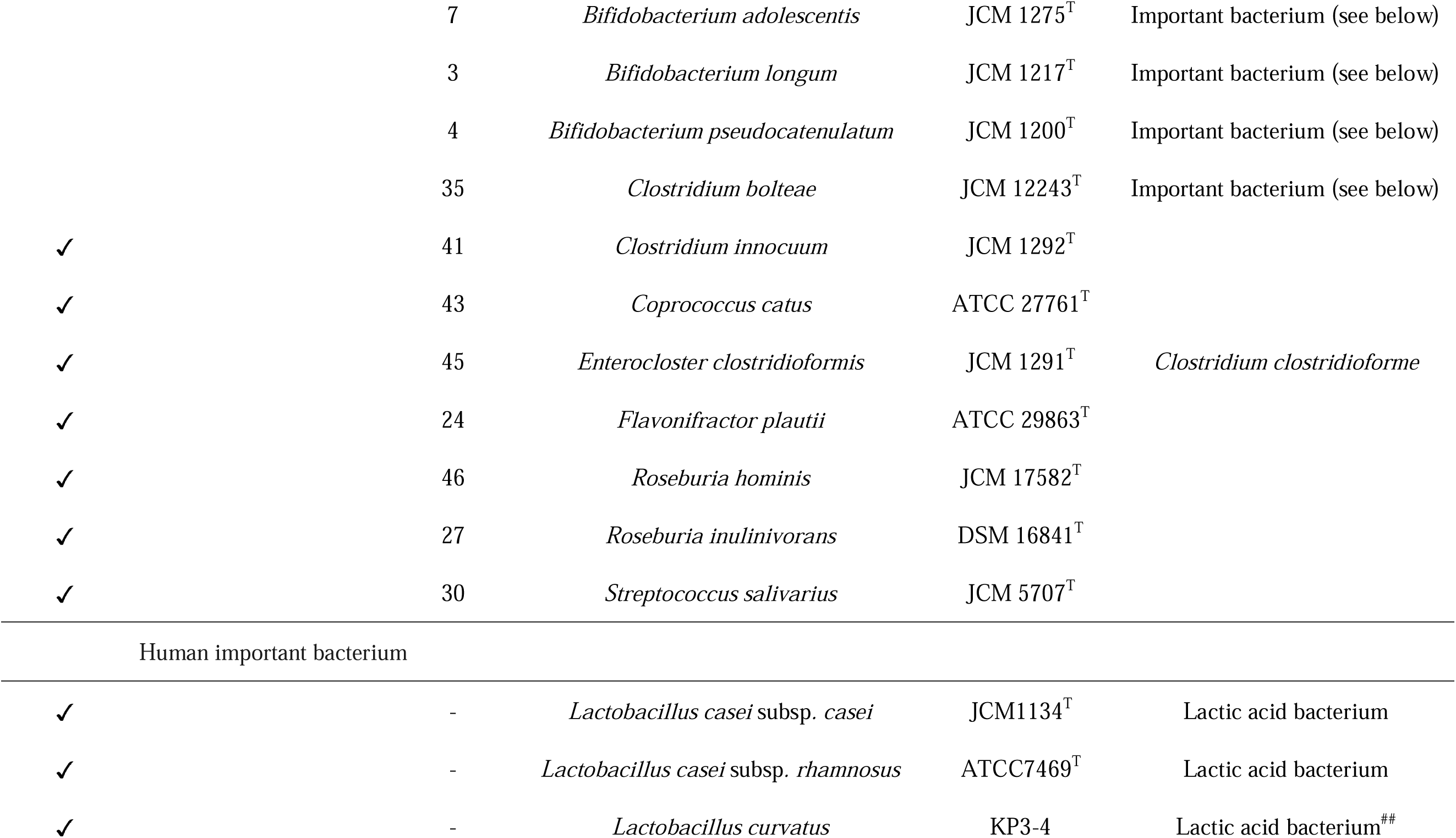
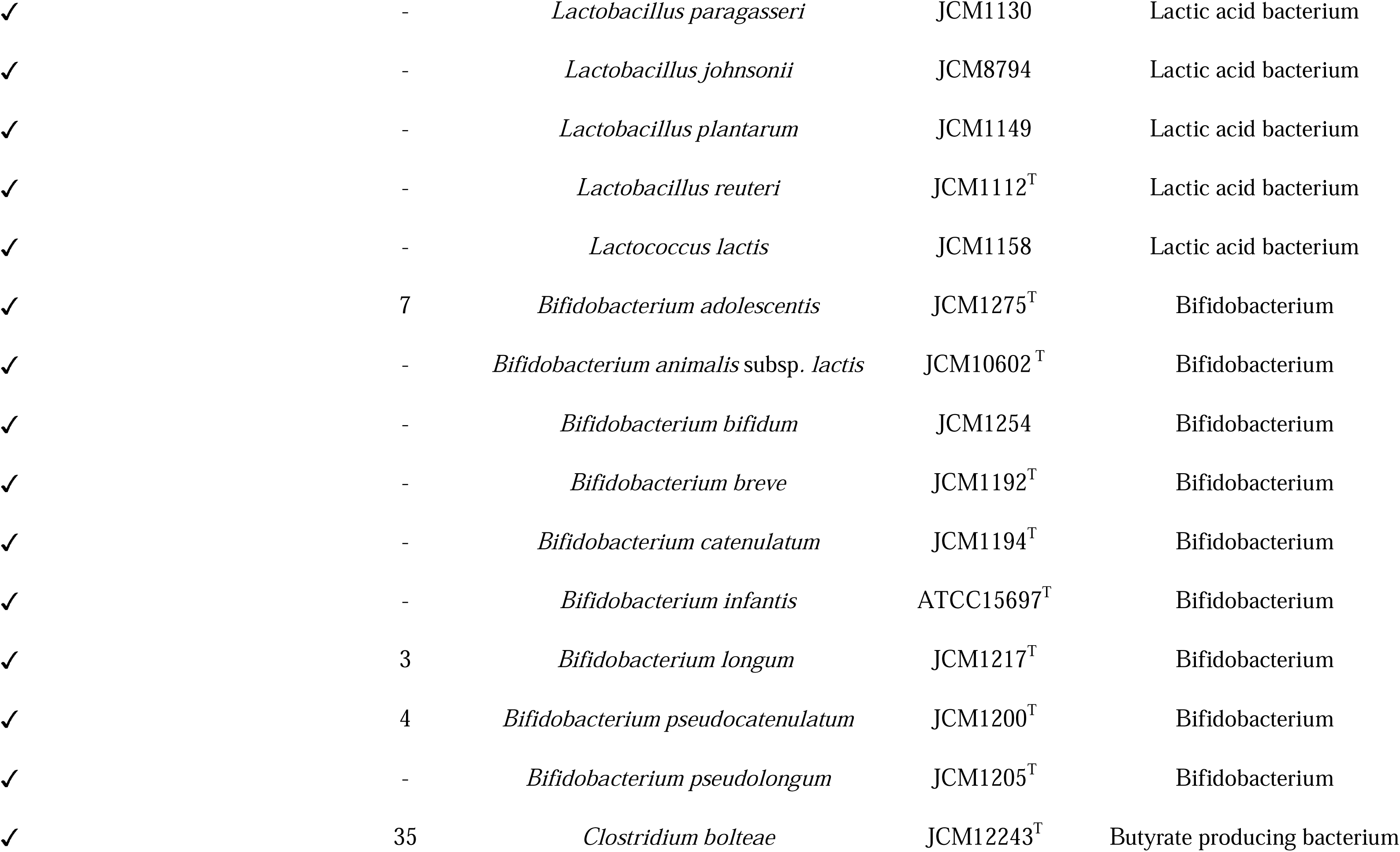
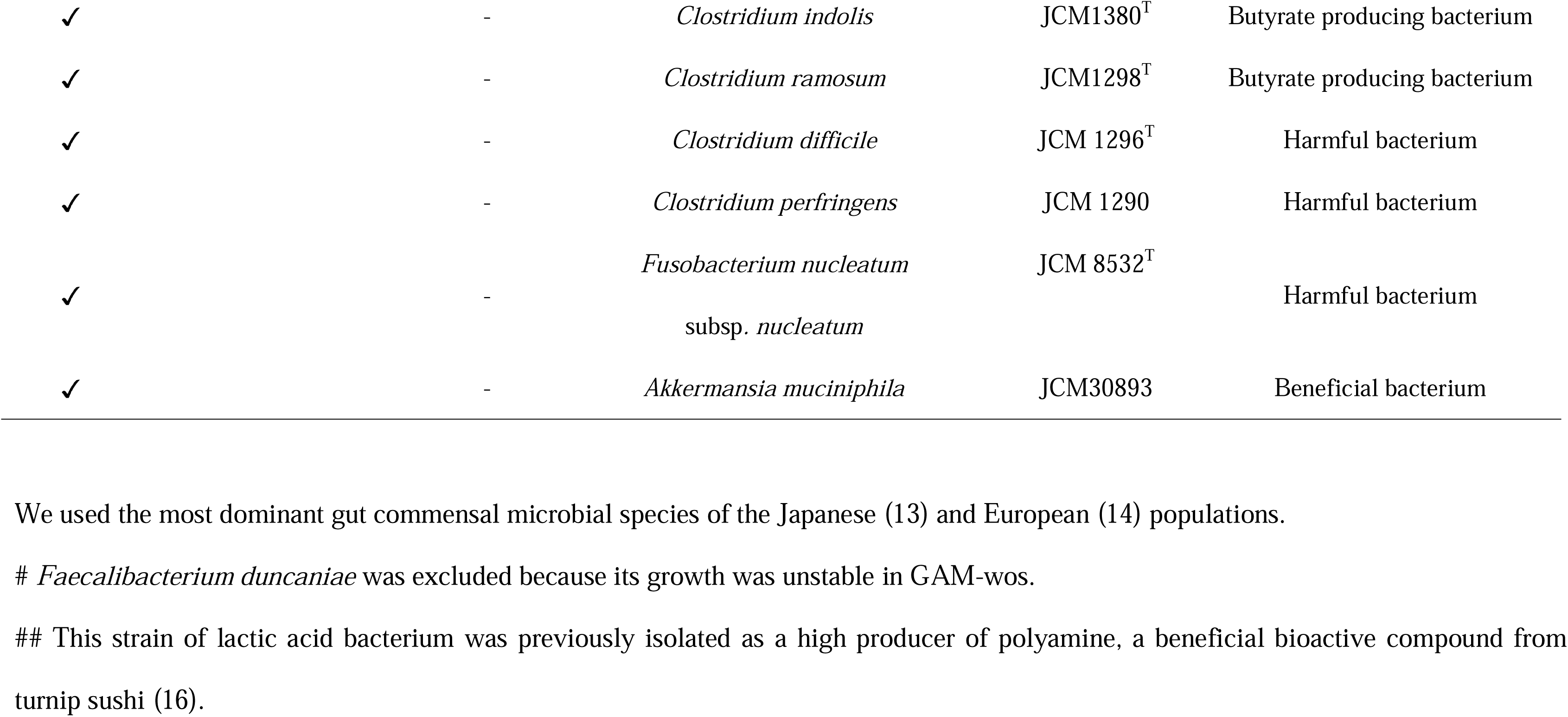
Details of dominant and important human gut bacteria used in this study.

Effect of the addition of 0.5% (w/v) GlcN to sugar-removed GAM medium on the growth of the most dominant species of human intestinal microbiota was observed (Supplementary Fig. 1). The addition of 0.5% GlcN to the medium promoted the growth of 42 of the 50 most dominant species of human intestinal microbiota (*Anaerostipes hadrus, Anaerotruncus colihominis, Bacteroides caccae, Bacteroides dorei, Bacteroides eggerthii, Bacteroides finegoldii, Bacteroides fragilis, Bacteroides intestinalis, Bacteroides ovatus, Bacteroides stercoris, Bacteroides thetaiotaomicron, Bacteroides uniformis, Bacteroides vulgatus, Bacteroides xylanisolvens, Blautia hansenii, Butyrivibrio crossotus, Clostridium asparagiforme, Clostridium clostridioforme, Clostridium innocuum, Clostridium nexile, Collinsella aerofaciens, Coprococcus comes, Dorea longicatena, Enterococcus faecalis, Eubacterium rectale, Eubacterium ventriosum, Faecalibacterium prausnitzii, Holdemania filformis, Parabacteroides johnsonii, Parabacteroides merdae, Pseudoflavonifractor capillosus, Roseburia hominis, Roseburia intestinalis, Roseburia inulinivorans, Ruminococcus gnavus, Ruminococcus lactaris, Ruminococcus obeum, Ruminococcus torques, Streptococcus salivarius, Subdoligranulum variabile, Bifidobacteorium adolescentis, Bifidobacterium longum, Clostridium bolteae*) [Supplementary Fig. 1]. The addition of 0.5% GlcN to the medium promoted the growth of 17 of the 24 important species (seven out of eight lactic acid bacteria species (*Lactobacillus casei subsp. casei, Lactobacillus casei subsp. rhamnosus, Lactobacillus curvatus, Lactobacillus paragasseri, Lactobacillus johnsonii, Lactobacillus plantarum, Lactococcus lactis*), 5 out of 9 bifidobacterial species (*Bifidobacterium bifidum, Bifidobacterium breve, Bifidobacterium infantis, Bifidobacterium longum, Bifidobacterium pseudocatenulatum*), 3 out of 3 butyric acid-producing bacteria species (*Clostridium bolteae, Clostridium indolis, Clostridium ramosum*), and 2 out of 3 harmful bacteria species (*Clostridium perfringens and Fusobacterium nucleatum subsp. nucleatum*) [Supplementary Fig. 2].

The growth data were substituted into the above formula to calculate the growth promotion and inhibition effects of GlcN on the bacterial species (Fig. 2). Of the total 70 bacterial species tested, the growth of 57 (81%) was statistically significantly promoted by 0.5% (w/v) GlcN. Especially, the growth of *A. colihominis, P. capillosus,* and *R. hominis* was promoted more than five-fold by 0.5% (w/v) GlcN (Fig. 2). In contrast, the growth of six species (9%; *Clostridium scindens, Coprococcus catus, Dorea formicigenerans, Lactobacillus reuteri, Bifidobacterium adolescentis and Clostridium difficile*) was statistically significantly suppressed by 0.5% (w/v) GlcN.

**Fig. 2.**
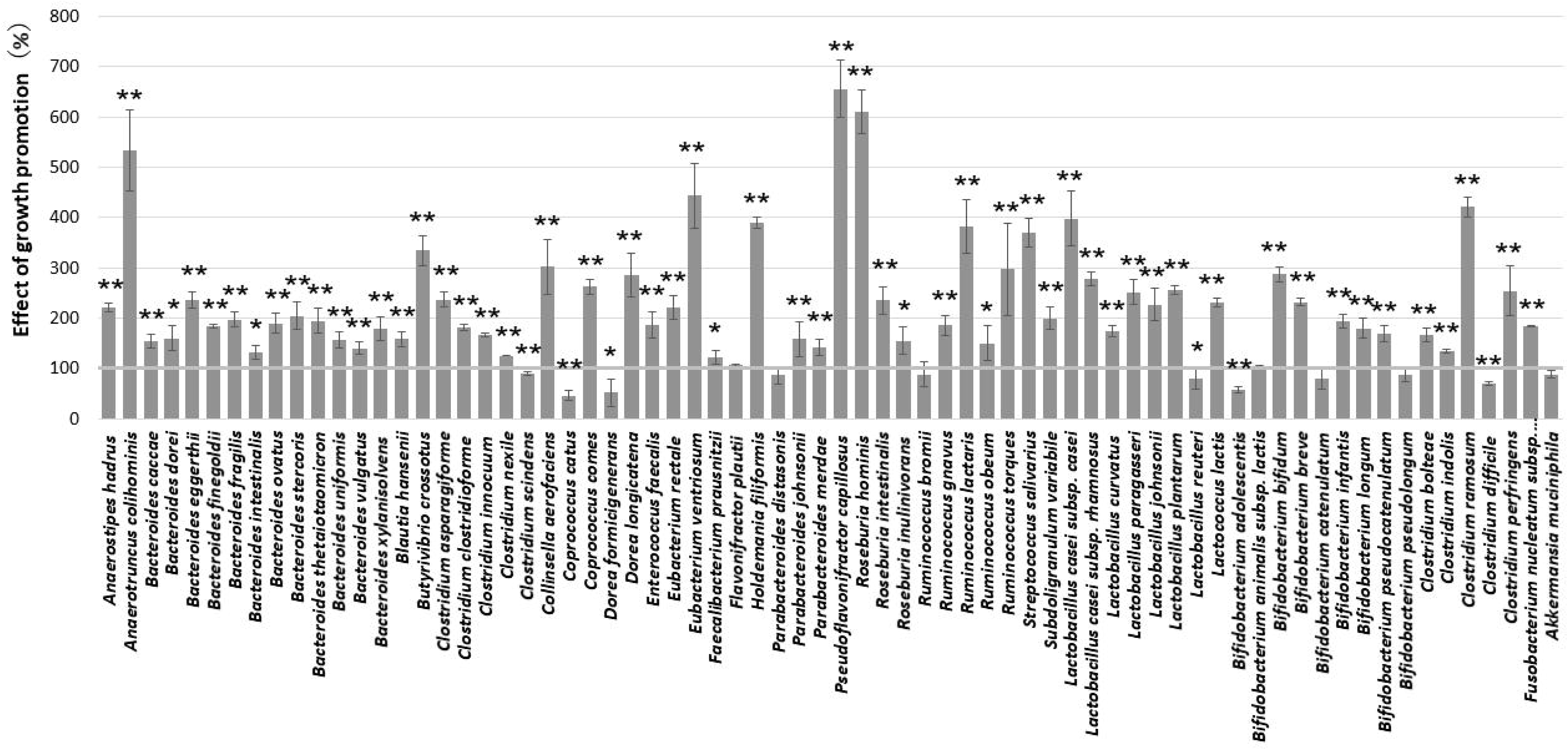
Evaluation of the effect of GlcN on growth of the 70 human gut important bacteria species. The growth of the most dominant and important intestinal bacteria in the intestinal microbiota, cultured in a sugar-free medium supplemented with GlcN, was used to calculate the growth promotion and inhibition effects of GlcN on intestinal bacterial species. Briefly, after 48 h of anaerobic incubation, growth was measured at an optical density of 600 nm (OD_600_). To compare the ability of the bacterial species to utilize GlcN, the OD_600_ ratio was obtained by dividing the bacterial growth (OD_600_) in the medium supplemented with GlcN by that without GlcN. All outcomes were presented as mean ± SD.

Next, we conducted an open-label, single-arm trial on bowel movements in 29 healthy individuals to determine the effects of the most common dose [22,23]. The bowel movement diary of the four defecation items (stool characteristics, color, odor intensity, and residual stool sensation) was assessed using the respective scales (Fig. 3).

**Fig. 3.**
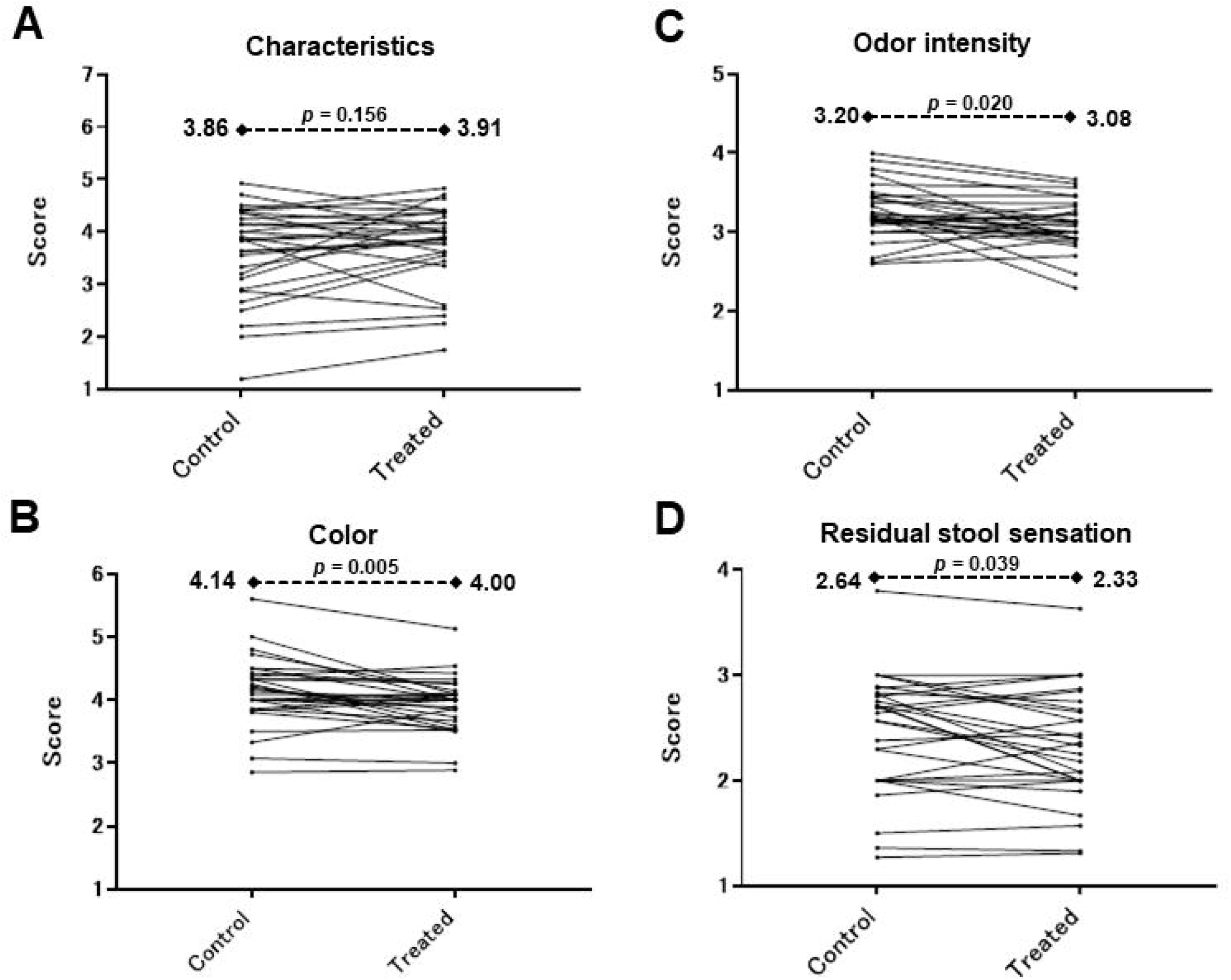
Individual influence of dietary GlcN on the four defecation items. Stool characteristics, color, odor, and residual stool sensation were assessed in 29 healthy participants. (A) Stool characteristic (Bristol stool scale) classified the stool form into 7 scales, from “Separate hard lumps, like nuts,” to “Watery, no solid pieces, entirely liquid.” Each answer was tallied. The closer it is to 4, the more likely the patient is to have a better bowel movement status. (B) Stool color was assessed on a 6-point scale (from 1 = yellow to 6 = dark brown to almost black), and the scores were tallied. (C) Stool odor intensity was assessed on a 5-point scale (1 = very weak, 5 = very strong), and the scores for each answer were tallied. (D) Residual stool sensation was assessed on a 4-point scale (1 = no sensation to 4 = full sensation). All outcomes were presented as median values. Statistical analysis was performed using the Wilcoxon signed-rank test before and during the 2 weeks of intake.

The individual effects of dietary GlcN on the four defecation items are shown in Figure 3. Stool characteristics were assessed using the Bristol Stool Scale. There was no difference between the stool characteristics during the 2-week pre-intake and during the GlcN-intake (3.60 ± 0.87 and 3.75 ± 0.76, respectively; *p* = 0.156) [Table 2]. The color of the stool was significantly different from that during the 2-week pre-intake and during the GlcN-intake (4.14±0.54 and 3.93±0.43, respectively; *p* = 0.005) [Table 2]. There was a significant difference in odor between the stools during the two periods (3.25 ± 0.34 and 3.09 ± 0.31, respectively; *p* = 0.020) [Table 2]. Residual stool sensation was significantly different between the two experimental periods (2.46±0.56 and 2.33±0.54, respectively; *p* = 0.039) [Table 2].

**Table 2.** Defecation items assessed on scales.

| Parameter | Pre | Post | *p* value |  |
| --- | --- | --- | --- | --- |
| Stool Characteristics | 3.60 ± 0.87 | 3.75 ± 0.76 | *p* = 0.156 |  |
| Stool Colors | 4.14 ± 0.54 | 3.93 ± 0.43 | *p* = 0.005 | ** |
| Stool Odor Intensity | 3.25 ± 0.34 | 3.09 ± 0.31 | *p* = 0.020 | * |
| Residual Stool Sensation | 2.46 ± 0.56 | 2.33 ± 0.54 | *p* = 0.039 | * |

All outcomes were presented as mean ± SD.
\*Statistical analysis was performed using the Wilcoxon signed-rank test before and during the 2 weeks of intake.
\* : $p < 0.05$ \*\* : $p < 0.01$ \*\*\* : $p < 0.001$

The individual effects of dietary GlcN on the four constipation parameters are shown in Figure 4. Numerical measurements of the four constipation items (amount of stool, frequency of defecation and flatulence, and days of defecation) were assessed in the 29 participants (Fig. 4). The stool amount was significantly different between that during the pre-intake 2-week period and during the intake of GlcN (9.3±5.0 and 13.6 ±7.3, respectively; *p* < 0.0001) [Table 3]. The frequency of defecation was significantly different between the two periods (4.05±1.26 and 5.59±1.55, respectively; *p* < 0.0001) [Table 3]. The frequency of farts tended to increase with GlcN intake, but the difference was not statistically significant (*p* = 0.059). The days of defecation were significantly different between the two experimental periods (3.69± 1.06 and 4.97±1.03; *p* < 0.0001) [Table 3].

**Fig. 4.**
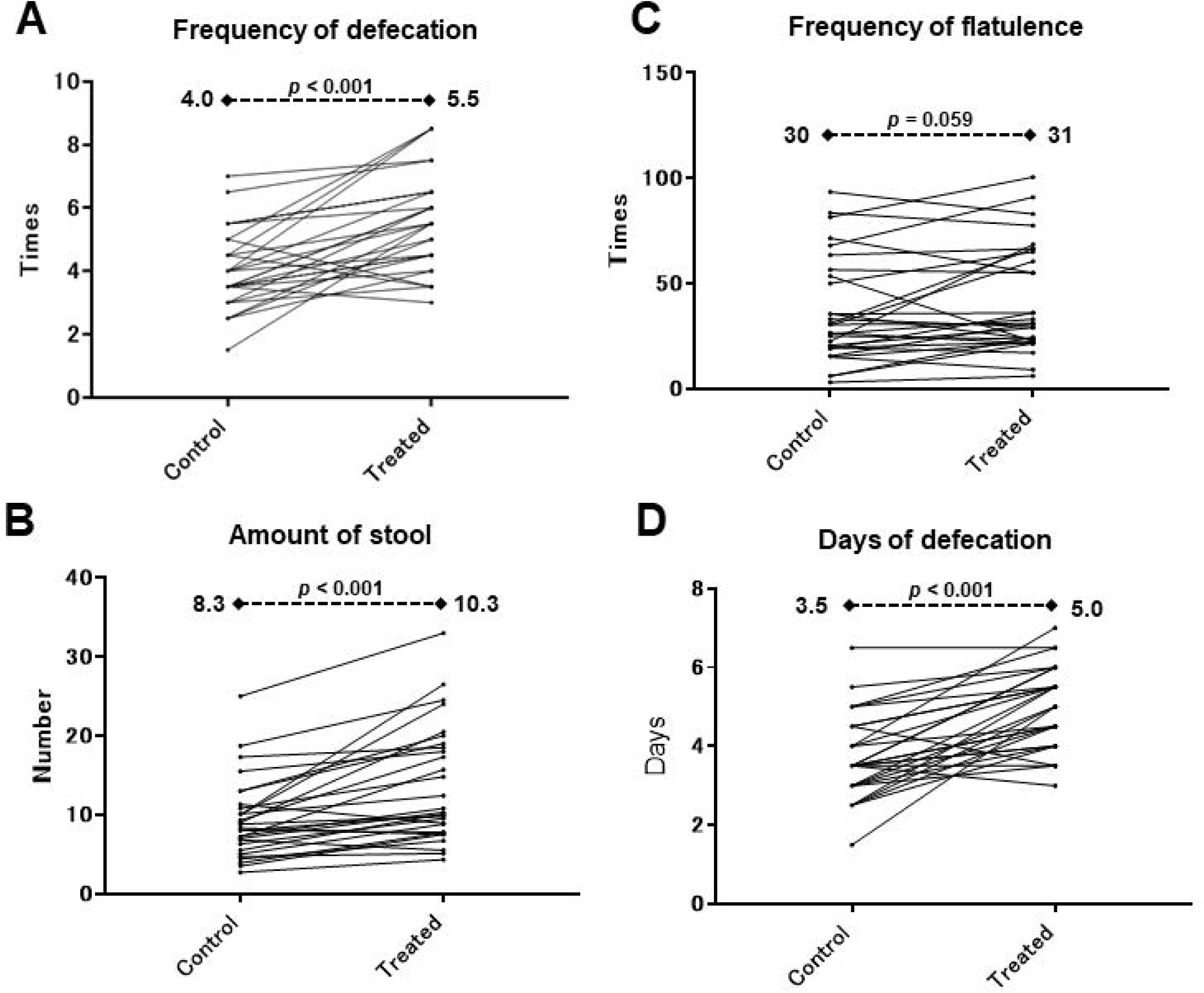
Individual influence of dietary GlcN on the four constipation items. The amount of stool, frequency of defecation and flatulence, and number of days of defecation were assessed in 29 healthy participants. (A) The amount of stool per a week was assessed as the estimated number (average of the 2 weeks) of large chicken eggs (large size) converted from the stool samples. (B) The frequency of defecation (average of the 2 weeks) was assessed as the number of defecation events per a week. (C) The frequency of flatulence was assessed as the number of (average of the 2 weeks) of flatulence events in the 2 weeks tested. (D) The number of days of defecation per a week was assessed as the number (average of the 2 weeks) of days of defecation events in 2 weeks. All outcomes were presented as median values. Statistical analysis was performed using the paired Student’s t-test before and during the 2 weeks of intake.

**Table 3.** Constipation items assessed by values.

| Parameter | Pre | Post | *p* value |  |
| --- | --- | --- | --- | --- |
| Amount of stool | 9.3 ± 5.0 | 13.6 ± 7.3 | *p* < 0.001 | *** |
| Frequency of defecation | 4.05 ± 1.26 | 5.59 ± 1.55 | *p* < 0.001 | *** |
| Frequency of flatulence | 36.4 ± 25.0 | 42.3 ± 25.8 | *p* = 0.059 |  |
| Number of days of<br>defecation | 3.69 ± 1.06 | 4.97 ± 1.03 | *p* < 0.001 | *** |

2 All outcomes were presented as mean ± SD.
\*Statistical analysis was performed using the paired Student's t-test before and during the 2 weeks of intake.
\* : $p < 0.05$ \*\* : $p < 0.01$ \*\*\* : $p < 0.001$

## DISCUSSION

We found that a wide range of tested gut bacteria utilized GlcN *in vitro*. Conventionally available prebiotics, such as raffinose, 1-kestose, lactulose, galacto-oligosaccharides, and fructo-oligosaccharides, have been reported to promote the growth of a wide range of gut bacteria using similar assay systems [17]. On the other hand, a new prebiotic disaccharide, D-galactosyl-β1→4-l-rhamnose, selectively enhanced the growth of Bifidobacterium and specifically suppressed the growth of the harmful bacterium *Clostridioides difficile* [17]. Thus, D-galactosyl-β1→4-l-rhamnose was thought to become a next generation prebiotic, in contrast to conventional prebiotics that promote the growth of a broad spectrum of gut bacteria. Considering the above, in this study GlcN appears to function similarly to that of conventional prebiotics and not like next generation prebiotics.

Among the 70 human gut bacteria tested in this study, the greatest growth activity (over 5-fold) was shown by *A. colihominis*, *P. capillosus*, and *R. hominis* (Fig. 2). *A. colihominis* is a butyrate producer that ameliorated experimental autoimmune encephalomyelitis, which is associated with the induction of regulatory T cells in the lymph, in a mouse model [24]. *P. capillosus* is a gram-negative, non-sporulating species belonging to the Clostridium cluster IV [25]. This cluster includes species that produce acetate and butyrate [26]. *R. hominis* is also a butyrate producer [27]. *R. hominis*, a host gut colonizer, shows upregulation of its bacterial genes involved in metabolism and motility; in addition, its colonization upregulates host genes related to antimicrobial peptides, gut barrier function, and Toll-like receptor signaling [28]. *R. hominis* was also recently reported to be depleted in stool samples of obese individuals compared to lean controls, and its abundance was negatively correlated with body mass index and serum triglycerides [29]. All three species are related to the production of SCFAs, such as butyrate and acetic acid.

A previous clinical report of 10 individuals showed that dietary 1500 mg GlcN and 1200 mg chondroitin increased the abundance of four *Lachnospiraceae* genera, two *Prevotellaceae* genera, and *Desulfovibrio*, compared to the placebo [30]. This study included chondroitin as a possible confounding factor in the test substance, making it difficult to interpret the effects of GlcN on gut bacteria. Another clinical report with a smaller sample size of 6 individuals showed that dietary 3000 mg GlcN, which was double the dose used in this trial, showed a trend towards reducing constipation with no significant increase in distinctive genera [12]. Thus, a few studies have explored the potential relationship between GlcN intake and changes in gut microbiota composition; however, the details of the effects of GlcN on the gut microbiota and bowel movements in humans are still unknown. To address the limitations of previous studies [12,30], we employed a single-arm design with 29 participants, providing a larger sample size than that of previously reported studies [12,30]. The increased number of participants may enhance the statistical power of the analysis, allowing for a more comprehensive assessment of the effect of supplementation with GlcN alone, at a normal dose of 1500 mg/day, on bowel movements.

In human trials, the Bristol scale score is associated with gut microbiota richness and composition [31]. In this study, the stool statistics evaluated using the Bristol scale did not change during GlcN intake (Fig. 3); therefore, the gut microbiota richness and composition might not change. However, it is possible that GlcN increases the number of total bacteria or specific bacteria without changing their richness. An increase in bacterial count in vitro may lead to an increase in fecal volume in clinical trials (Fig. 4). In addition, the significant increase in SCFA-producing bacteria observed in vitro may have led to pH-mediated stool color changes [32]. Taken together, the results of this study suggest a potential increase in butyrate-producing bacteria and improved SCFA-mediated bowel movements following GlcN consumption. However, this study has certain limitations. It did not measure the metabolites contained in the culture medium or feces; therefore, future research is needed to measure the production of SCFAs, which have a significant effect on pH [32]. In addition, as the in vitro experiments in this study involved culturing intestinal bacteria alone, the interactions between the intestinal bacteria themselves or with the host were not considered. Additionally, in clinical trials, a blinded randomized study should be conducted to accurately evaluate the results and eliminate placebo effects [33].

In conclusion, we revealed that dietary GlcN could stimulate the growth of a wide range of gut microbiota components in vitro, including beneficial butyrate-producing bacteria. Additionally, to our knowledge, we provide the first clinical data showing that a small dose of GlcN alone favorably influences colon health. GlcN significantly improved defecation and constipation. This could be attributed to the effects of GlcN on improving bowel movements, potentially mediated by the metabolites of the gut microbiota, indicating a possible probiotic-like effect.

## Data Availability

All data produced in the present study are available upon reasonable request to the authors.

## Abbreviations

GAM: Gifu anaerobic medium;
GB: medium,
GAM: and blood medium;
GlcN: Glucosamine;
SCFA: short-chain fatty acids;
wos: without sugar;
ATCC: American Type Culture Collection;
DSMZ: German Collection of Microorganisms and Cell Cultures GmbH
JCM: Japan Collection of Microorganisms

## ACKNOWLEDGMENTS

The microbiome strains were provided by the American Type Culture Collection (ATCC), German Collection of Microorganisms and Cell Cultures GmbH (DSMZ), and Japan Collection of Microorganisms (JCM). This research was partially supported by JSPS KAKENHI (to S.K.). This research was also partially funded by the Toyo Institute of Food Technology (a nonprofit organization) [to T.S. and S.K.].

## AUTHOR CONTRIBUTIONS

T. S. and S. K. designed the study. T. S. wrote the manuscript. S.K. reviewed and edited the manuscript. S.S., Y.A., and K.S. conducted in vitro experiments. T.S. conducted the clinical trials. H.S. supervised the clinical trial. All the authors have read and approved the final version of this manuscript.

---

**Supplementary Fig. 1.**
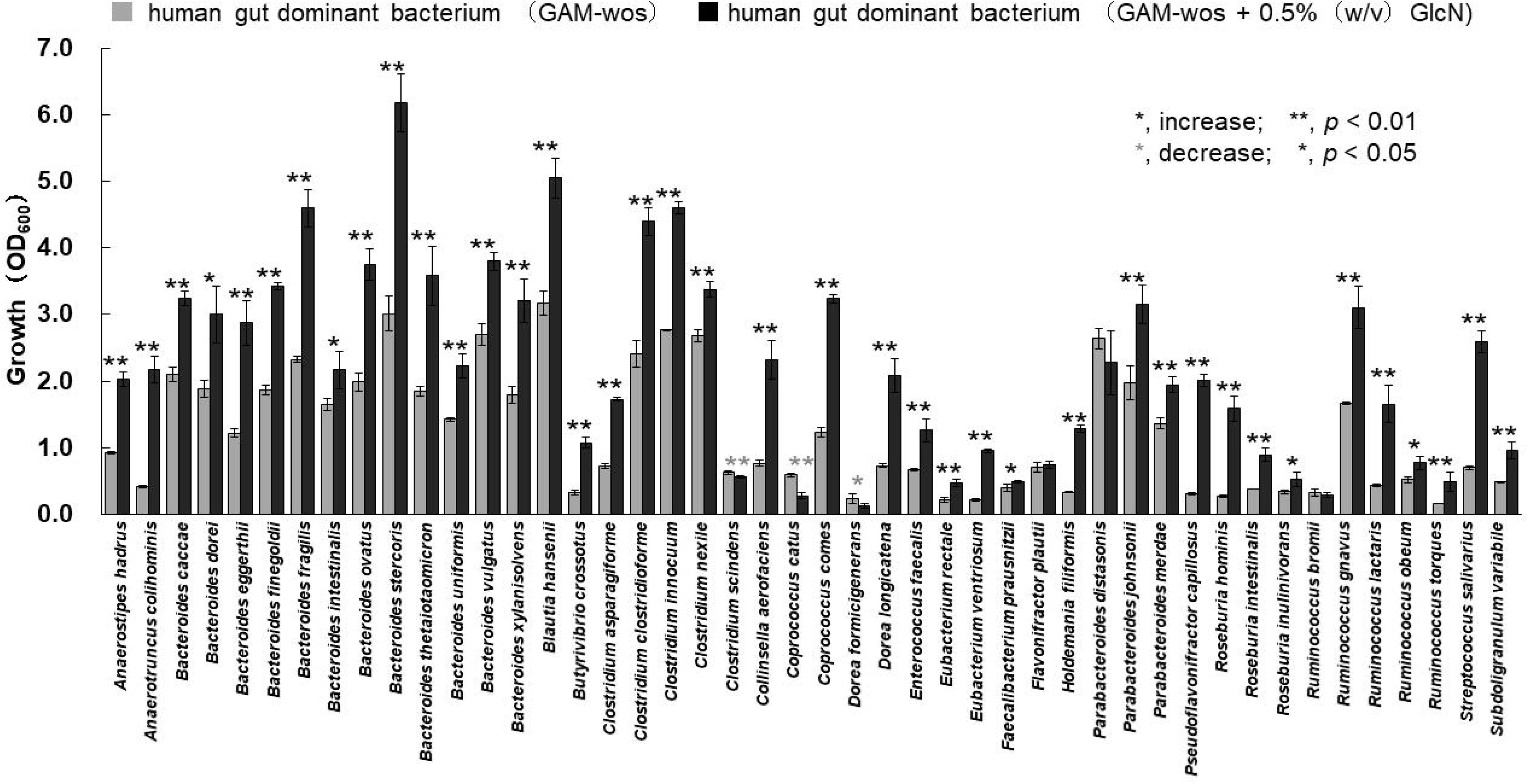
Growth of the human gut-dominant bacteria species with the addition of GlcN. The growth of the 46 most dominant human gut bacteria in the intestinal microbiota, which were cultured in sugar-free medium supplemented with 0.5% GlcN, were substituted to calculate the growth promotion and inhibition effects of GlcN on intestinal bacterial species. After 48 h of anaerobic incubation, growth was measured at an optical density of 600 nm (OD_600_).

**Supplementary Fig. 2.**
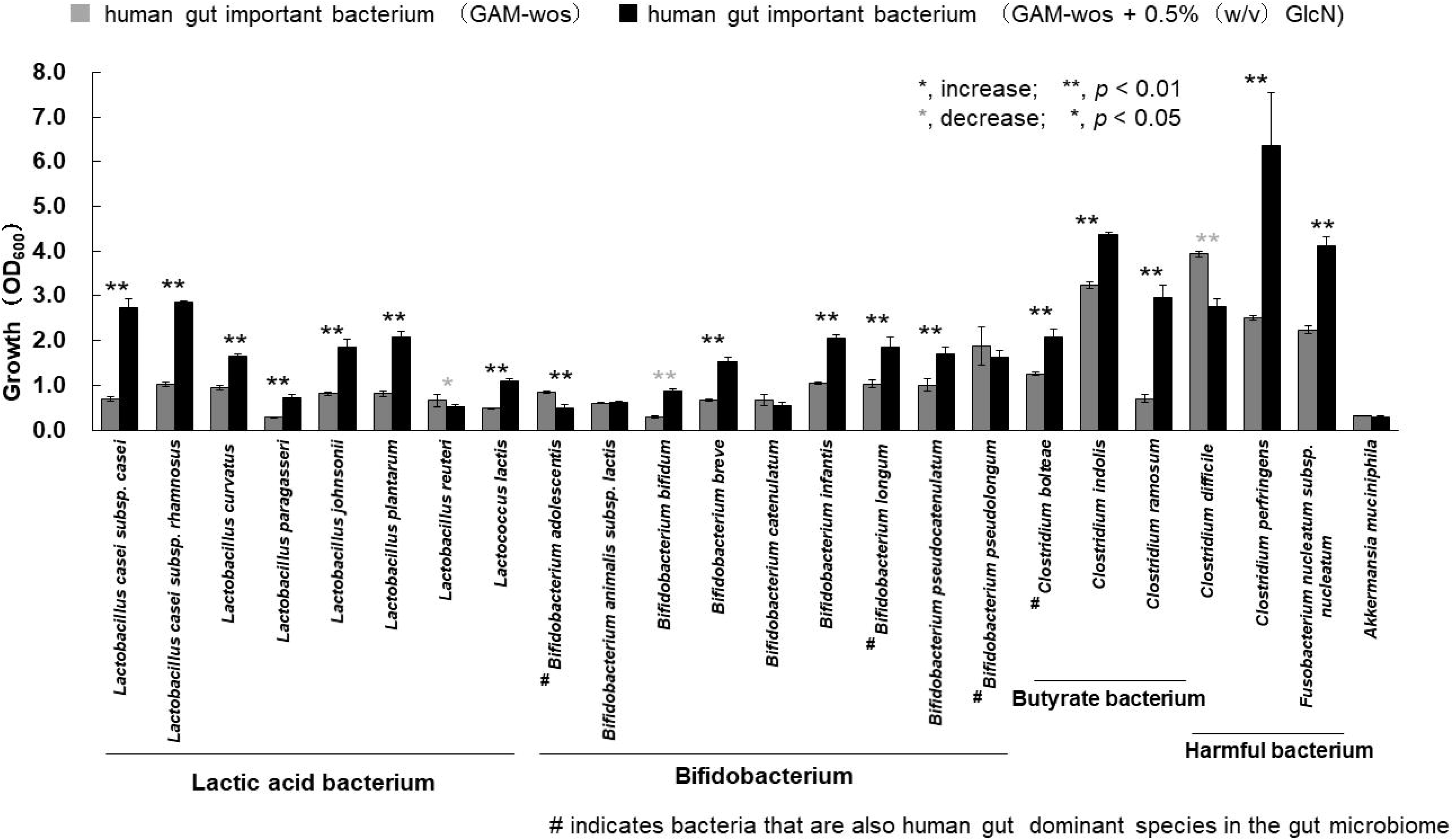
Growth of the human gut-important bacteria species with the addition of GlcN. The growth of the 24 important human gut bacteria in the intestinal microbiota, which were cultured in sugar-free medium supplemented with 0.5% GlcN, were substituted to calculate the growth promotion and inhibition effects of GlcN on intestinal bacterial species. After 48 h of anaerobic incubation, growth was measured at an optical density of 600 nm (OD_600_).

